# Evidence-Based WASHmobile Anticipatory Action Framework to Reduce Cholera Outbreaks during Extreme Weather Events in the Democratic Republic of the Congo

**DOI:** 10.64898/2026.10.22.26366235

**Authors:** Kelly Endres, Presence Sanvura, Jean-Claude Bisimwa, Alves Namunesha, Willy Felicien, Lucien Bisimwa, Alain Mwishingo, Camille Williams, Jamie Perin, Justin Bengehya, Ghislain Maheshe, Jean-Claude Kulondwa, Cirhuza Cikomola, Anna Dimitrova, Christine Marie George

## Abstract

Extreme weather contributes to cholera outbreaks globally. We examined associations between extreme weather and cholera through 4 years of surveillance at 115 healthcare facilities in the Democratic Republic of the Congo, and built an anticipatory action framework through WASHmobile for water, sanitation, and hygiene mobile messaging during extreme weather. Drought and wet conditions were defined using 3-month Standardized Precipitation Indices with CHIRPS data. A test-negative design assessed associations between extreme weather and culture-confirmed *Vibrio cholerae* among 1,068 diarrhea patients. Odds of cholera were higher during drought (OR: 1.94, 95% CI: 1.17-3.20) and extreme wet conditions (OR: 2.22, 95% CI: 1.39-3.55). During drought conditions in our randomized controlled trial of 2334 participants, handwashing with soap significantly declined in the standard arm but not the WASHmobile arm. Findings informed an anticipatory action framework where pre-defined climate triggers activate WASHmobile as an early warning system to reduce cholera outbreaks during extreme weather.

## INTRODUCTION

Extreme weather events, such as droughts and floods, are increasing globally and contributing to cholera outbreaks, which have reached historical highs (1). Low- and middle-income countries (LMICs) are expected to experience disproportionate health impacts of extreme weather due to vulnerable health systems and water and sanitation infrastructure (2). Cholera remains a major public health concern, with an estimated 1.3-4 million annual cases globally (3). Understanding how climate variability influences cholera transmission is increasingly important for outbreak prediction, preparedness, and prevention. This need is urgent given the increase in extreme weather events, such as the current strong El Niño, with elevated risks of floods, drought, and extreme heat (4). In Africa, El Niño is strongly associated with increased cholera incidence and severity (5).

However, most studies of climate and diarrheal disease rely on all-cause diarrhea or aggregated case counts, which cannot distinguish cholera from other enteric pathogens (6, 7).

Water, sanitation, and hygiene (WASH) interventions are critical for preventing cholera and other diarrhea outbreaks. However, evidence on WASH intervention effectiveness under extreme weather is limited (8, 9), with literature focusing mostly on seasonal effects (10–12). In a meta-analysis, WASH interventions reduced diarrhea risk by 33% during wet seasons, versus 18% during dry seasons (10). Studies are needed that investigate WASH intervention effectiveness during extreme weather events and that design climate-resilient WASH interventions that can be activated before these events.

Early warning systems can reduce diarrhea outbreaks resulting from extreme weather, but only if linked to timely interventions. Anticipatory action formalizes this link by developing triggers, actions, and resources that can be mobilized before a hazard’s impacts occur (13). Mobile health (mHealth) interventions are promising for promoting climate-resilient WASH behaviors, given their scalability and ability to deliver targeted, behavior-specific messaging at high-risk times. WASHmobile is an mHealth program that uses SMS, voice, and interactive voice response (IVR) messages to promote water treatment and hygiene behaviors among populations at high risk of diarrhea outbreaks (14). Previous WASHmobile randomized controlled trials (RCTs) in Democratic Republic of the Congo (DRC) and Bangladesh reduced healthcare facility visits for diarrhea, diarrhea prevalence, and stunting in children under 5 years, with sustained WASH behavior improvements (15, 16).

This study addresses these evidence gaps by linking culture-confirmed *Vibrio cholerae* among admitted diarrhea patients to climatic conditions and by measuring WASH behaviors during extreme weather within an RCT. We aimed to 1) evaluate how climatic conditions are associated with cholera and WASH behaviors in a humanitarian context in the DRC; 2) investigate through an RCT WASHmobile effectiveness on WASH behaviors during these climatic conditions; and 3) translate these findings into an evidence-based anticipatory action framework to reduce cholera outbreaks linking climate triggers to climate-resilient WASHmobile messages to guide future responses to extreme weather events.

## METHODS

### Study Design

This study was conducted in urban Bukavu, South Kivu Province, eastern DRC. Passive cholera surveillance was conducted at 115 healthcare facilities between March 2020 and September 2024 in collaboration with the DRC Ministry of Health, recruiting diarrhea patients and their household members. Within this surveillance, a two-arm cluster-randomized RCT of 2626 participants of the WASHmobile PICHA7 program was completed between December 2021 and December 2023 with household as the cluster unit (ClinicalTrials.gov: <u>NCT05166850</u>). The Catholic University of Bukavu (7107) and the Johns Hopkins Bloomberg School of Public Health Institutional Review Board (9848) approved the study protocol. Written informed consent was obtained from all adult participants, written consent from a parent or guardian for participants <18 years of age, and assent from children aged 12-17 years.

Patients admitted to the healthcare facility with diarrhea (three or more loose stools within 24 hours) had stool tested for *V. cholerae* by bacterial culture per our published methods (17). Cholera patients were defined as diarrhea patients with a *V. cholerae*-positive sample.

The detailed trial protocol is published elsewhere (15). Participants were assigned to one of two arms and followed for 12 months: a standard message arm, receiving standard DRC healthcare facility discharge guidance on oral rehydration solution use, or the WASHmobile PICHA7 arm, receiving this guidance plus weekly WASH voice, IVR quiz, and SMS messages (full protocol in Appendix File 1).

### WASH Outcomes

RCT outcomes were (1) observed handwashing with soap at stool- and food-related events (assessed by 5-hour structured observation) and (2) *E. coli* and free chlorine in stored drinking water (assessed by unannounced spot checks). Evaluation activities were conducted at Day 7 and Months 1, 3, 6, 9, and 12. WHO drinking water guidelines of <1 colony-forming unit (CFU)/100 mL of *E. coli* and of >0.2 mg/L free chlorine were used as cutoffs (18).

### Climate Exposures

GPS coordinates of all diarrhea patient households were paired to climate data. Precipitation data were obtained from Climate Hazards Group InfraRed Precipitation with Station data (CHIRPS) at 0.05° (∼5.5 km) resolution (19).

Climate exposures were selected from previous literature to capture rainfall and drought conditions associated with diarrhea risk (6–8, 20–33). Drought was characterized using the Standardized Precipitation Index (SPI) (34), calculated using CHIRPS from 1990-2024 using the SPEI R package (35). For each day, 3-month accumulated precipitation was fitted to a gamma distribution over a 1990-2024 reference period and transformed to a standard normal distribution. The 3-month timescale was selected to capture cumulative medium-term moisture conditions biologically relevant for drought impacts on diarrhea outbreaks, consistent with previous analyses (36). SPI-3 categories were extreme drought (≤ −2.00), severe drought (-1.5 to -1.99), moderate drought (-1.0 to -1.49), near normal (-0.99 to 0.99), moderate wet (1.0 to 1.49) severe wet (1.5 to 1.99), and extreme wet (≥2.00) per standard definitions (34). Precipitation anomalies in the previous 4 weeks were used to assess shorter-term rainfall deviations, defined as standard deviations from the location- and study period-specific long-term mean precipitation over the study period (6, 22).

### Statistical Analyses

A test-negative design comparing culture-confirmed cholera patients with culture-negative diarrhea patients was used to investigate associations between SPI-3 and cholera using logistic regression. Models were unadjusted and run for all ages and stratified by age (<5 and ≥5 years), given higher diarrhea severity in young children (37). Vaccination status, gender, and age were included in sensitivity analyses.

Associations between SPI-3 and RCT outcomes were investigated using generalized estimating equations with exchangeable correlation to account for household clustering. WASH outcomes were handwashing with soap at least once during a structured observation, *E. coli* >1 CFU/mL and free chlorine >0.2 mg/L in source and stored drinking water. Water quality models were adjusted for water source type (improved vs. unimproved per WHO Joint Monitoring Program guidelines (38)). WASH behavior were adjusted for socioeconomic status (SES) indicators (household income source (yes/no), people per sleeping room, and concrete floor (yes/no)). Study arm was also investigated as a potential effect modifier of the relationship between climatic exposures and WASH behaviors models. Baseline WASHmobile PICHA7 observations were excluded.

SPI was entered as a single categorical variable with near normal as the reference. Given previous studies finding increased diarrhea when intense rainfall occurred after dry periods (29, 39), 4-week precipitation anomaly (continuous) was investigated as a potential effect modifier of the relationship between drought SPI categories and cholera. All analyses were conducted using R (version 4.5.2).

### Anticipatory Action Framework Development

These findings informed an anticipatory action framework where pre-defined climate triggers would activate WASHmobile as an early warning system for cholera outbreaks during extreme weather, integrating climate data with surveillance to strengthen cholera prevention.

## RESULTS

### Cholera and Climate Exposures

Over the 4-year study period, 1068 diarrhea patients were captured across the 115 surveillance healthcare facilities. Among these diarrhea patients, 31% (326/1068) were culture confirmed cholera patients, including 11% (55/520) of children under 5 and 49% (271/548) of those 5 or older (Appendix Table (AT) 1). Mean age was 14.9 years (standard deviation: 18.6, range: 0-82.3), and 49% (528/1068) were female. Mean monthly rainfall was 109.68 mm/month (range: 9.09-280.00) (Figure 1).

**Figure 1.**
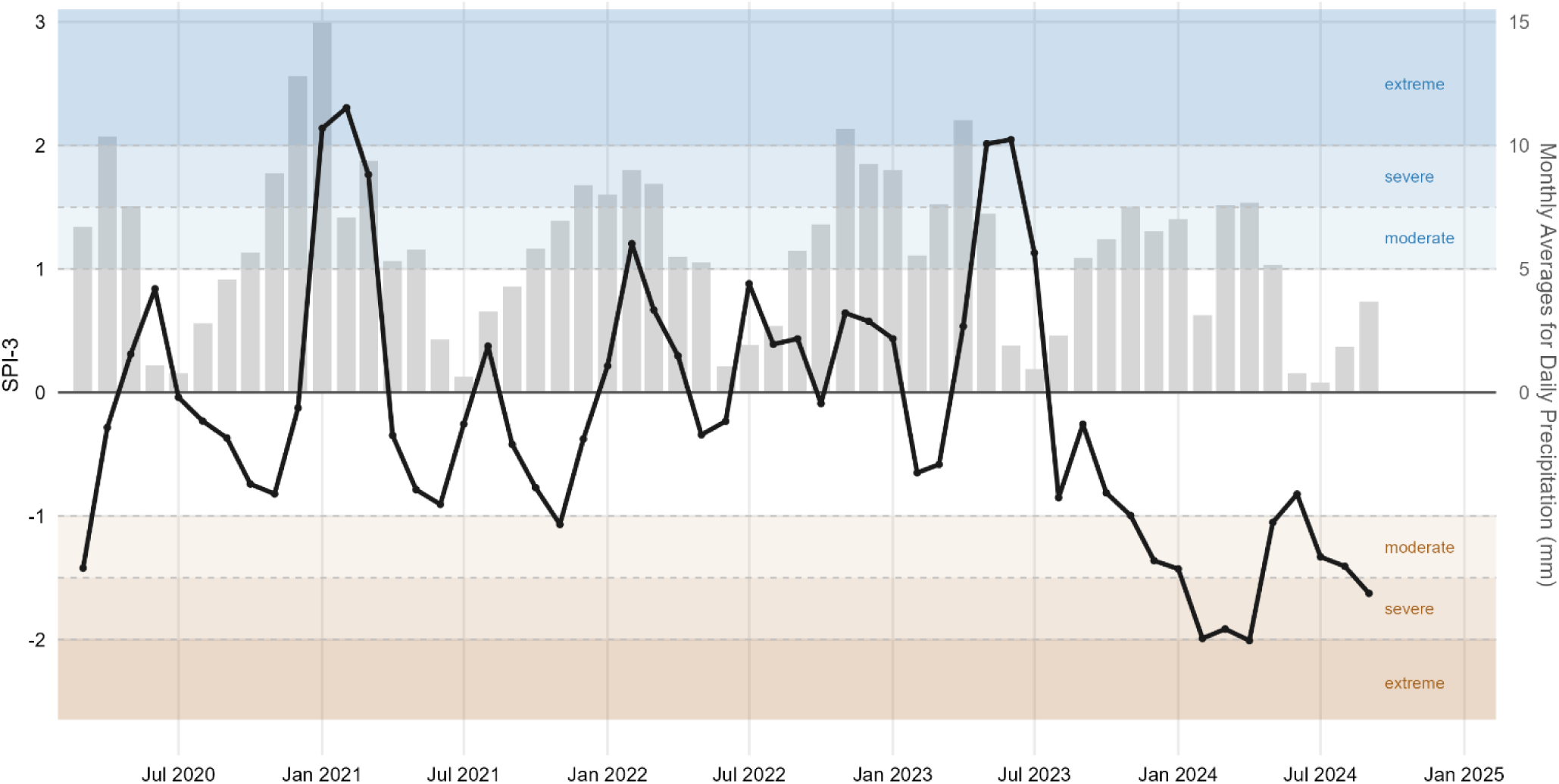
SPI-3 and monthly precipitation in Bukavu, South Kivu, DRC, March 2020 - September 2024. Bars show monthly precipitation (month-averaged daily maximum precipitation) from CHIRPS, and the black line shows SPI-3.

Among all ages, the odds of cholera were significantly higher during extreme wet conditions (odds ratio (OR): 2.22, 95% confidence interval (CI): 1.39-3.55) and severe wet (OR: 1.87, CI: 1.02-3.40) compared to near normal (Figure 2). Odds of cholera were also significantly higher during moderate drought among all ages (OR: 1.94, CI: 1.17-3.20), and children under 5 (OR: 3.11, CI: 1.17-8.31). The 4-week precipitation anomaly did not modify the association between moderate drought and cholera (AT 2A).

**Figure 2.**
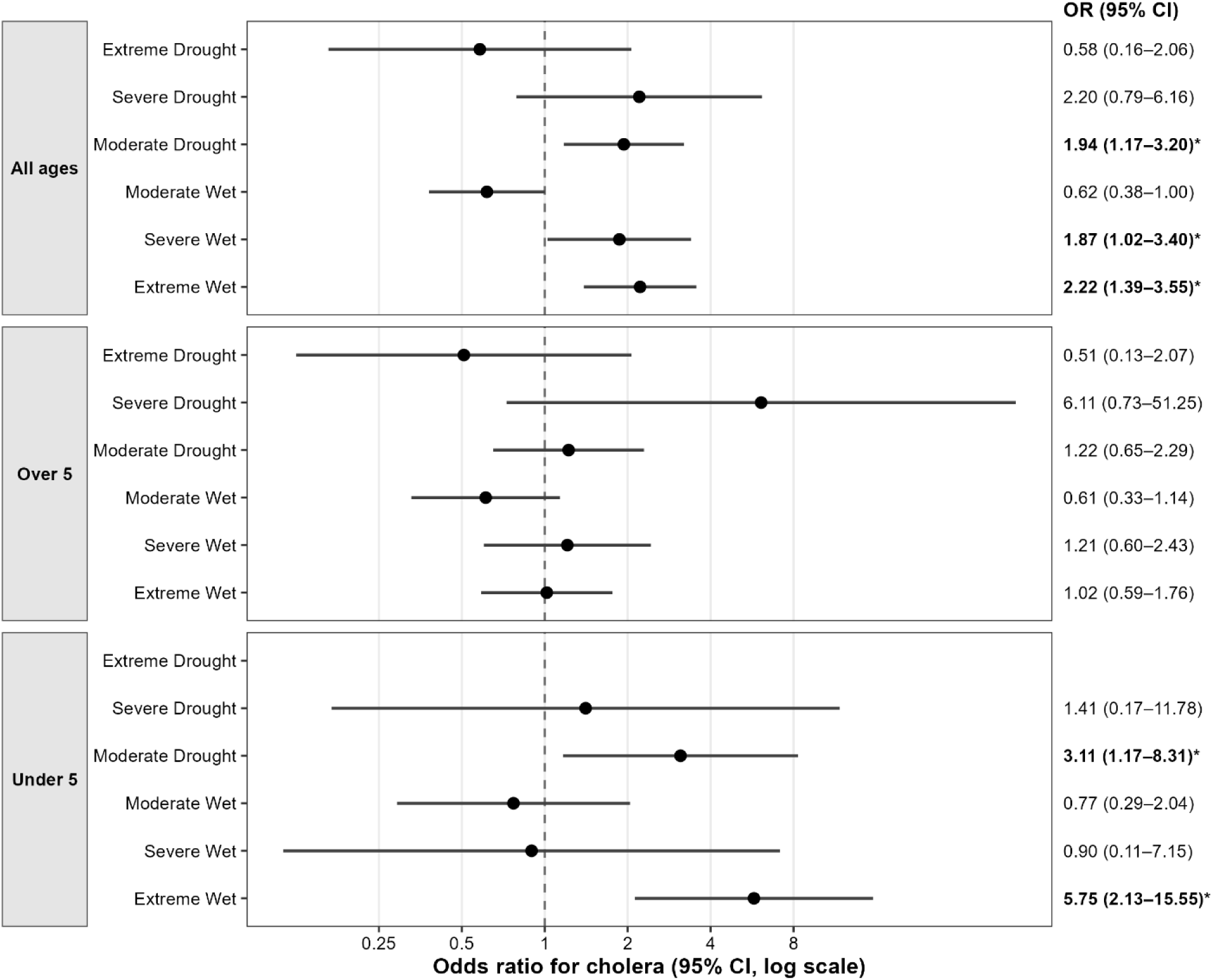
Associations between SPI-3 categories and cholera among admitted diarrhea patients, by age group (N = 1068). Odds ratios (ORs) and 95% Wald confidence intervals (CIs) are from logistic regression models using a test-negative design, with culture-negative diarrhea patients as the comparison group. Models were fitted separately for all ages (N = 1068), ≥5 years (N = 548), and <5 years (N = 520). SPI-3 ORs are from a single model per age group with near normal as the reference. Circles denote point estimates. The dashed vertical line marks OR = 1, and the x-axis is on a log scale. Asterisks indicate p < 0.05. Cholera refers to diarrhea patients that were found to have *V. cholerae* in stool by bacterial culture. Extreme drought were excluded for children under 5 years due to sparse observations. Extreme drought not calculated for under 5 due to sample size constraints.

Sensitivity analyses adjusting for vaccination status and gender found no impactful change in results (AT 2B, 3B). After adjustment for age in years, associations among all ages remained significant for moderate drought and extreme wet conditions (AT 3B).

### Handwashing RCT Findings

Participants washing hands with soap at least once during structured observations was 7% (113/1566) in the standard arm and 47% (858/1831) in the WASHmobile PICHA7 arm (AT 1). During moderate drought conditions, the odds of handwashing were 84% lower than near normal in the standard arm (CI: 0.03-0.81) (1% moderate drought vs. 7% near normal) with no significant difference in the WASHmobile PICHA7 arm (OR: 1.44, CI: 0.81-2.55) (Figure 3, AT 4B) (57% vs. 45%). Consistent with this, we observed significant interaction between moderate drought conditions and the study arm on handwashing with soap prevalence (interaction OR = 8.45, 95% CI: 1.62-44.18; multiplicative interaction p-value = 0.011) (Figure 4, AT 4).

**Figure 3.**
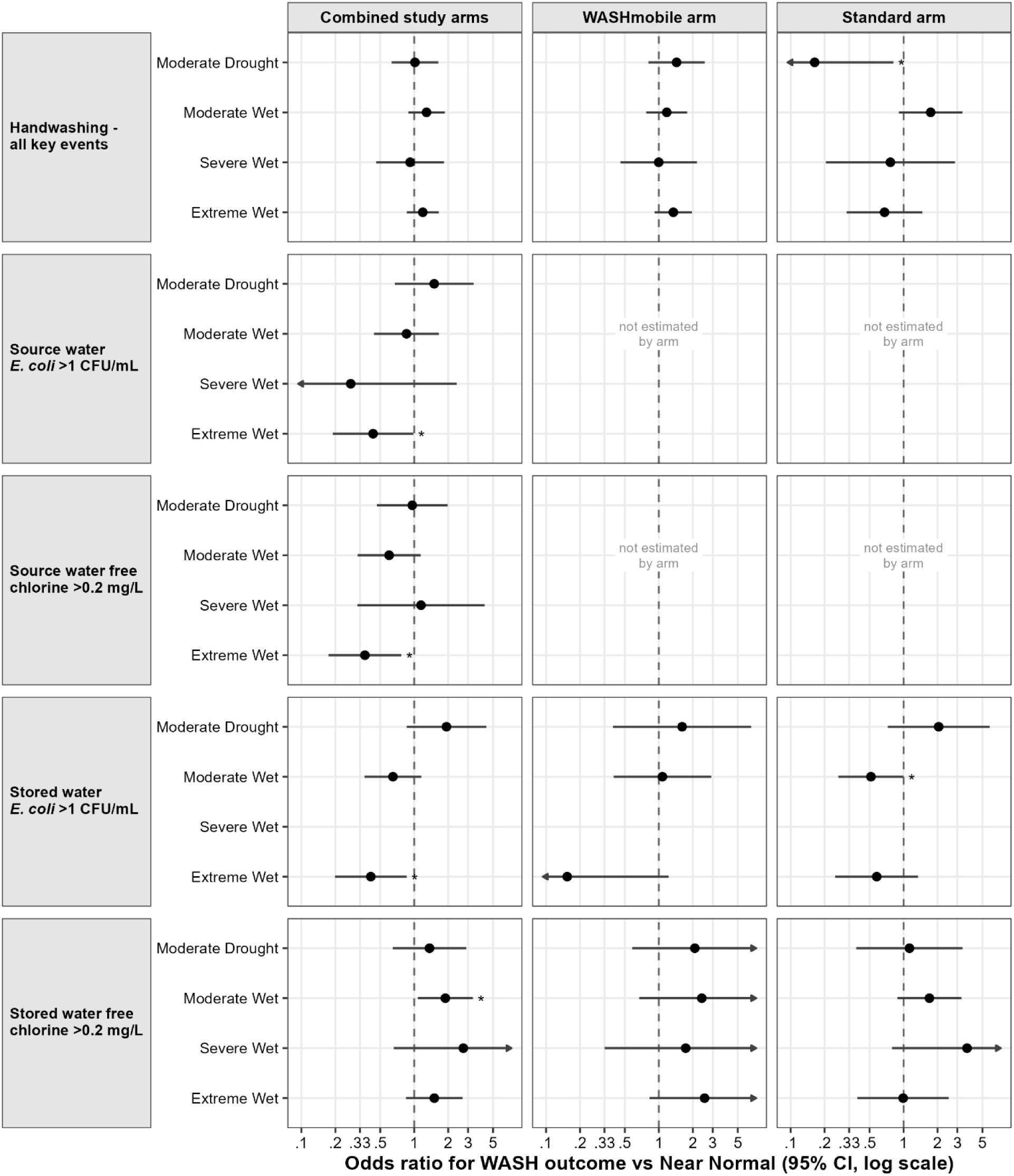
Associations between SPI-3 categories and WASH outcomes, overall and by study arm. Odds Ratio (OR) and 95% Wald Confidence Intervals (CIs) compare each SPI-3 category with near normal (reference). Estimates are from generalized estimating equation (GEE) models with an exchangeable correlation structure to account for household clustering. Water quality models are adjusted for water source type (improved vs. unimproved per WHO JMP guidelines (38)). All models except source water are adjusted for socioeconomic status (household income source, people per sleeping room, and concrete floor), and combined study arm models are adjusted for study arm. Severe and extreme drought were excluded due to sparse observations. Source water is unrelated to the intervention, so results were not stratified by study arm. Circles denote point estimates. Source water models were estimated for combined study arms only. Arrows indicate CIs extending beyond the plotted range. The dashed vertical line marks OR = 1, and the x-axis is on a log scale. Asterisks indicate p < 0.05. Estimates, p-values, and sample sizes are given in Appendix Table 3A. Observations with incomplete socioeconomic data excluded.

**Figure 4.**
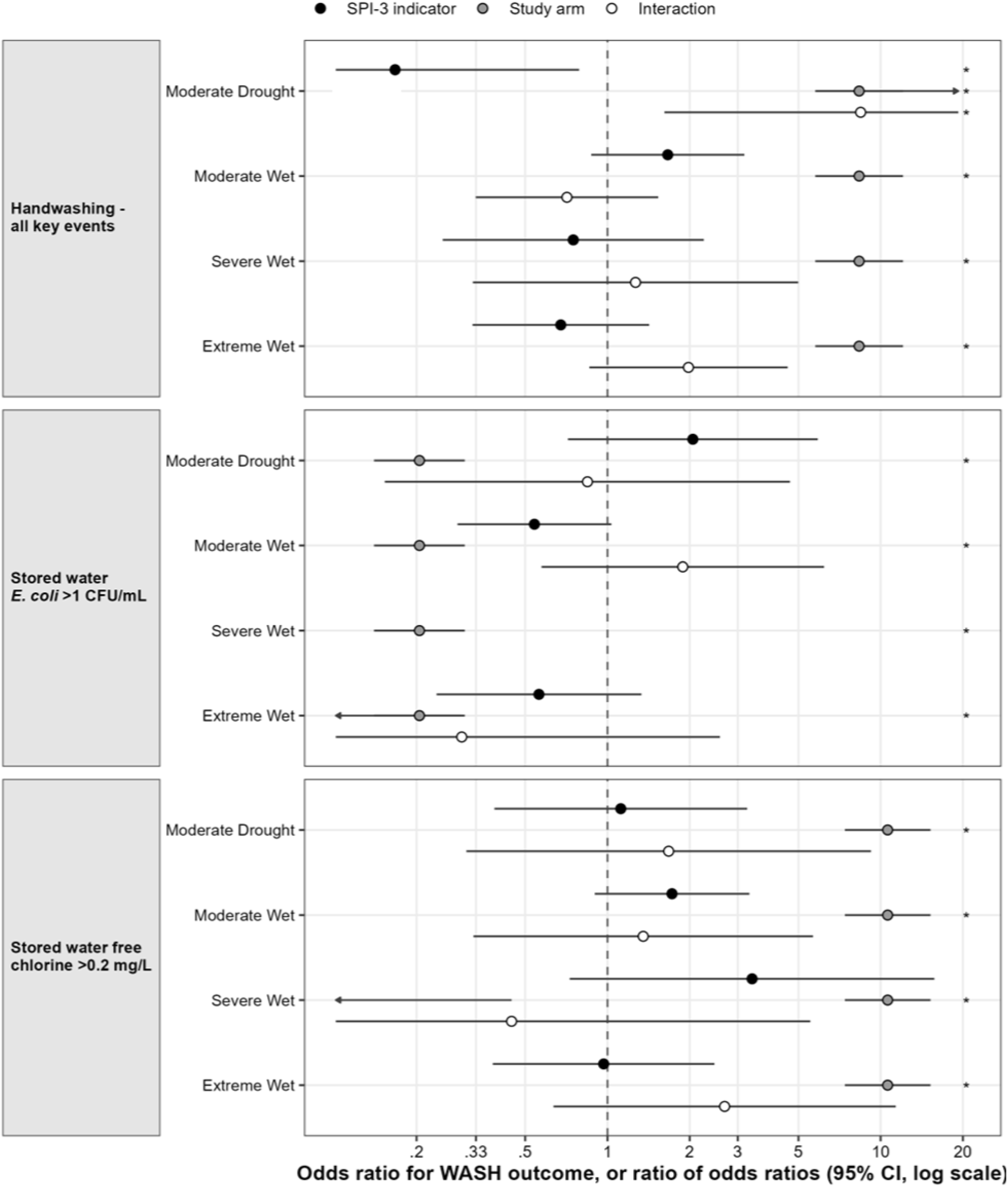
Effect modification of associations between SPI-3 categories and WASH outcomes by study arm. Points show three terms from one GEE model per outcome: the SPI-3 category Odds Ratio (OR) relative to near normal in the standard arm (filled circle), the study arm OR at near normal (grey circle; one estimate per model, repeated across category rows), and their interaction, a ratio of ORs (open circle), where >1 indicates the category’s OR is higher in the WASHmobile PICHA7 arm than in the standard arm. Covariates are as in Supplemental Table 4. Circles denote point estimates. Bars show 95% Wald Confidence Intervals (CIs), and arrows indicate CIs extending beyond the plotted range. The dashed vertical line marks a ratio of 1, and the x-axis is on a log scale shared across panels. Asterisks indicate p < 0.05. Estimates, together with the SPI-3 and study arm terms from the same models, are given in Appendix Table 4.

### Source Water Quality RCT Findings

For source water samples, 27% (214/796) had *E. coli* >1 CFU/mL and 33% (281/853) had free chlorine > 0.2 mg/L (AT 1). During extreme wet conditions, the odds of source water *E. coli* >1 CFU/mL were 57% lower than near normal (CI: 0.19-0.98; 28% vs. 15%) and 63% lower for free chlorine >0.2 mg/L (CI: 0.17-0.77; 3% vs. 17%) (Figure 3, AT 3A, AT 1).

### Stored Drinking Water Quality RCT Findings

Stored water *E. coli* >1 CFU/mL was found in 40% (237/591) of standard arm and 12% (58/502) of WASHmobile PICHA7 arm samples (AT 1). The odds of stored water *E. coli* were significantly lower during moderate wet than near-normal conditions in the standard arm (OR: 0.51, CI: 0.26-0.99; 29% near normal vs. 42% moderate wet), but not the WASHmobile PICHA7 arm (OR: 1.08, CI: 0.40-2.91; 12% vs. 13%) (Figure 3, AT 1, AT 3A).

Stored water free chlorine >0.2 mg/L was found in 82% (441/538) of WASHmobile PICHA7 and 30% (187/629) of standard arm samples (AT 1). For both arms combined, the odds of stored water free chlorine >0.2 mg/L were 89% higher during moderate wet conditions compared to near normal (CI: 1.08-3.31; 62% moderate wet vs. 51% near normal) (AT 1, AT 3A) with no difference observed by study arm.

### Translating Findings into an Anticipatory Action Framework for WASHmobile Early Warning Systems to Reduce Cholera Outbreaks

Findings informed the design of an evidence-based anticipatory action framework where pre-defined climate triggers inform deployment of the WASHmobile early warning system to reduce cholera outbreaks (Figure 5). Triggers can be calculated from openly available CHIRPS data and through weather forecasting systems. The framework has three phases. First, seasonal climate forecasts, such as El Niño, activate readiness actions months in advance such as preparing WASHmobile message content, establishing partnerships with mobile network operators for message delivery, and securing chlorine tablet for e-voucher program and oral rehydration solution (ORS) stockpiles with local shops and pharmacies, and enhancing diarrhea patient surveillance at healthcare facilities. Second, CHIRPS, local rainfall records, and weather forecasting systems trigger early actions when risk thresholds are crossed. Since SPI-3 reflects cumulative precipitation, it evolves gradually and can be tracked as thresholds approach, providing lead time. Short-range weather forecasting systems such as the AI-enhanced WeatherNext3 add further early warnings, and the related WeatherNext 3’s cyclones model generates forecasts of tropical cyclone tracks, intensity, and wind (40). For example, Google’s Flood Hub provides free, daily river flood forecasts up to 7 days ahead, and urban flood forecasts up to 24 hours ahead, which can prompt flood-specific WASHmobile messages and chlorine e-voucher delivery in flood forecasted areas (41). Cyclone forecasts could similarly trigger WASHmobile messages and chlorine e-voucher delivery ahead of landfall in cyclone-prone settings. However, future studies are needed to test these messaging interventions, since this was not investigated in the current study. Third, when healthcare facility cholera surveillance identifies a threshold of suspected cholera cases, this case-trigger intensifies mobile messaging in affected areas through WASHmobile messaging and chlorine e-vouchers, as in our recent study (42).

**Figure 5.**
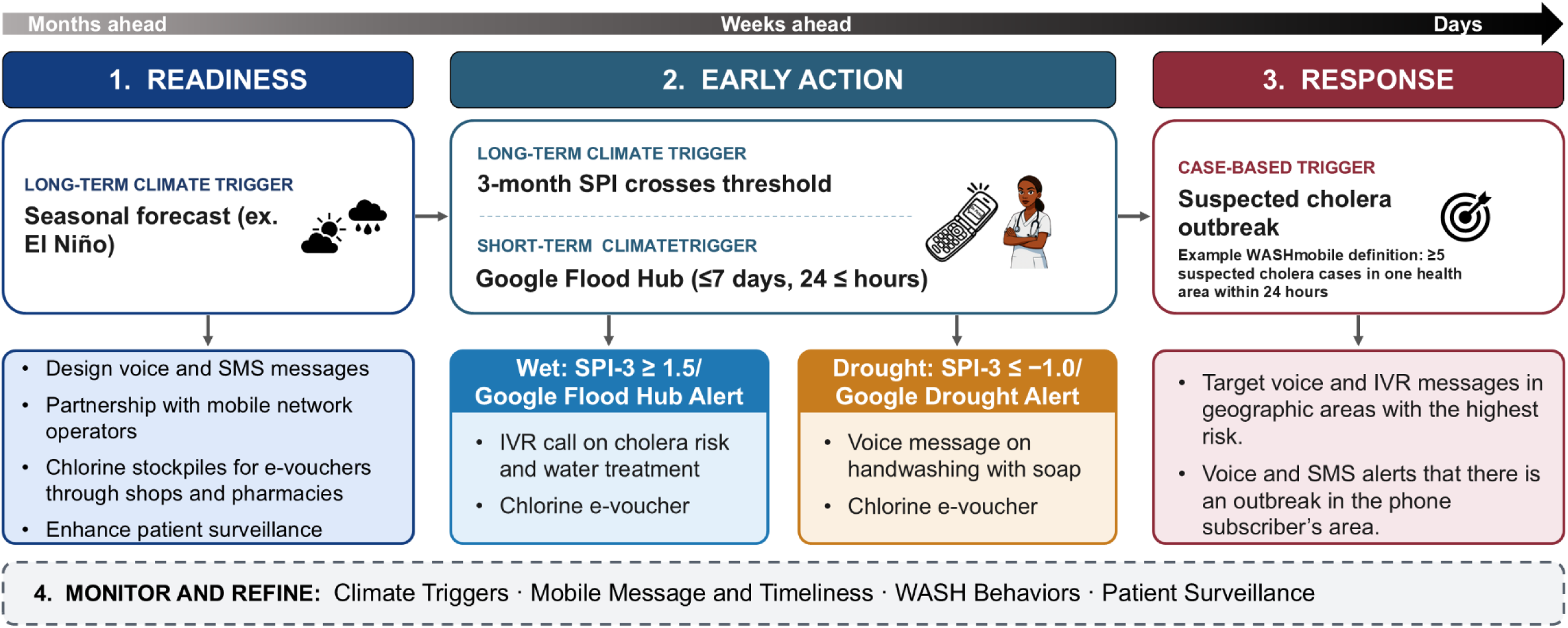
WASHmobile anticipatory action framework for reducing cholera outbreaks during extreme weather events. Monitoring and refinement spans all phases; IVR: interactive voice response; SPI: Standardized Precipitation Index.

Anticipatory actions are aligned with each study finding. Because extreme and severe wet conditions were associated with higher odds of cholera among diarrhea patients, extreme wet-conditions trigger activation of an IVR call to high-risk areas explaining the increased risk of cholera and correct chlorine tablet use and providing an e-voucher by SMS for free chlorine tablets at a local shop or pharmacy (Figure 6) (42). In addition, because moderate drought was associated with reduced handwashing with soap in the standard arm but not the WASHmobile PICHA7 arm, the drought trigger activates a voice message explaining the increased risk of cholera and that acknowledges water scarcity while emphasizes handwashing with soap as less costly than a diarrhea-related hospital visit. Alternative water sources (e.g., lake or river water) are also recommended for water for handwashing ( not drinking). These actions address the underlying mechanism rather than repeating generic hygiene reminders. Finally, we observed increased cholera patients during extreme wet and drought conditions, case-based trigger of a culture confirmed cholera case in a set geographic area (e.g. health area) activates delivery of a voice call explaining that cholera has been found in the phone subscribers neighborhood and the need for handwashing with soap and water treatment to prevent themselves and their family from getting cholera. This case-based trigger could also be based on the number of suspected cholera cases which we have defined as 5 suspected cholera cases in our recent study (42).

**Figure 6.**
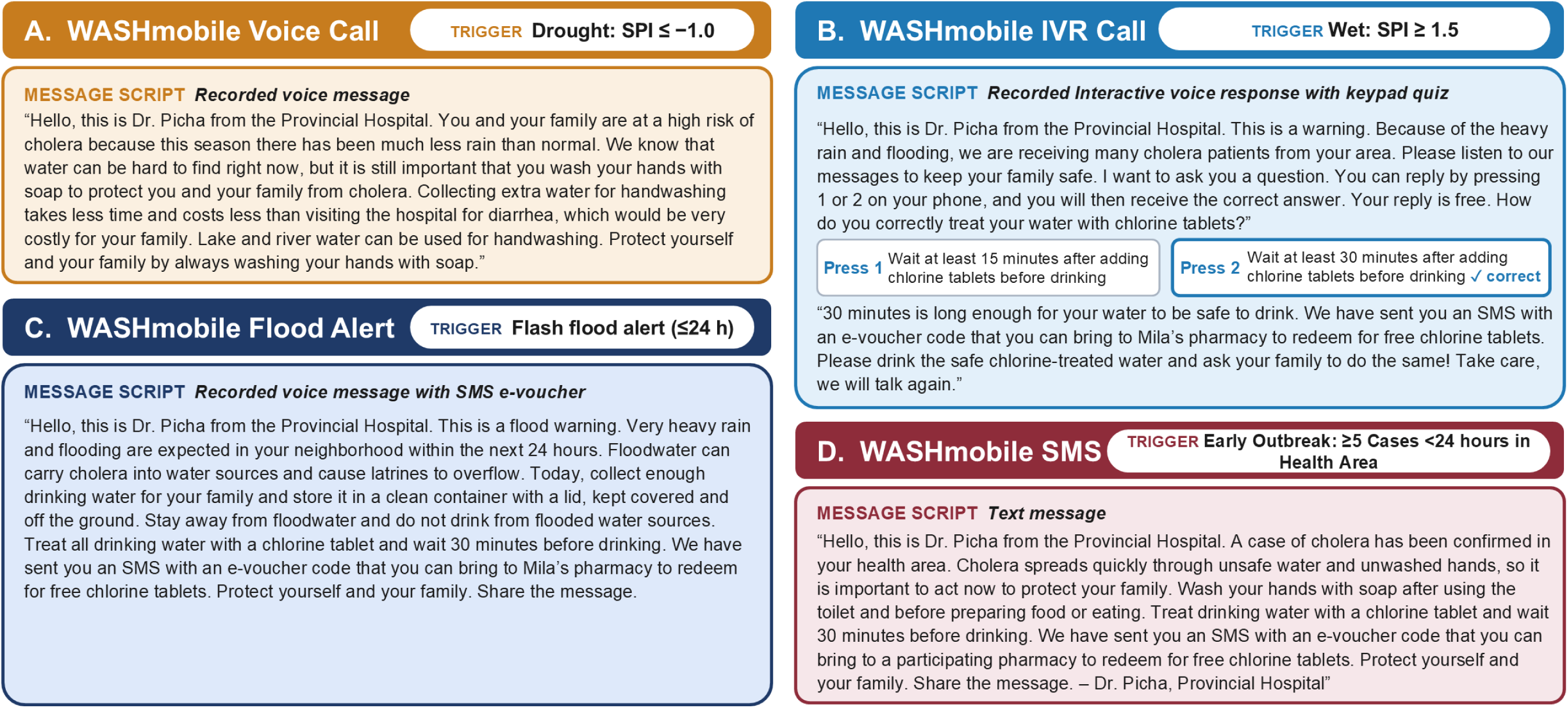
Example climate-resilient WASHmobile messages deployed as early actions under the WASHmobile anticipatory action framework. (A) Recorded voice call deployed during drought (SPI-3 ≤ −1.0). (B) Recorded interactive voice response (IVR) call deployed during severe or extreme wet conditions (SPI-3 ≥ 1.5). (C) Recorded voice flood alert with an SMS e-voucher, deployed when Google Flood Hub forecasts an urban flash flood within 24 hours. (D) Text message (SMS) with an e- voucher, deployed as a case-based response when ≥5 suspected cholera cases are identified in a health area within 24 hours. The drought message addresses water scarcity as a barrier to handwashing. The wet-condition messages promote household water treatment, and the IVR call includes a quiz on correct chlorine tablet use and an e-voucher for free chlorine tablets. The flood alert promotes storing covered drinking water before flooding, avoiding flooded water sources, and treating water afterwards. Scripts are shown in English for illustration.

Unlike traditional WASH interventions, which may be slower to adapt and scale, WASHmobile content can be revised and deployed within hours of a climate trigger at low marginal cost per message, reducing the cost of action. This is especially valuable in humanitarian settings such as eastern DRC, where health systems may lack capacity for rapid climate responses but mobile phone coverage enables low-cost, scalable communication.

## DISCUSSION

This study examined associations between extreme weather, cholera, and WASH behaviors in a humanitarian context in the DRC. The odds of cholera was significantly higher during moderate drought, extreme wet, and severe wet conditions. During moderate drought, handwashing with soap declined in the standard arm but not in the WASHmobile arm, suggesting WASHmobile sustained hygiene behaviors during drought conditions with limited water availability. We translated these findings into an anticipatory action framework where pre-defined climate triggers activate WASHmobile as an early warning system to reduce cholera outbreaks during extreme weather.

Extreme and severe wet conditions were associated with twice the odds of cholera among diarrhea patients of all ages compared to normal conditions. Potential mechanisms include flooding, surface runoff, and sanitation system overflow which increase environmental spread of *V. cholerae* and contaminate drinking water sources (43). Our test-negative design asked whether climatic conditions change the probability that a patient with diarrhea has cholera rather than another diarrheal pathogen. Higher *V. cholerae* positivity among admitted diarrhea patients could therefore reflect an increase in cholera, a decrease in other pathogen-specific types of diarrhea (e.g. rotavirus), or both.

Our findings therefore suggest prolonged extreme wet conditions may shift the etiologic composition of diarrhea for admitted patients toward *V. cholerae*, rather than simply increasing overall diarrheal disease burden, as often reported for rainy seasons (43). This is consistent with a meta-analysis finding that bacterial diarrheal pathogens are more common during rainy seasons, whereas viral and parasitic diarrhea showed weaker or more heterogeneous seasonal patterns (39).

Moderate drought was also associated with significantly higher odds of cholera. Potential mechanisms include reduced water availability, increased reliance on unsafe or unimproved water sources, higher pathogen concentration in depleted water sources (6), and reduced handwashing with soap due to water scarcity (44), the latter being supported by reduced standard arm handwashing during moderate drought. Wang et al.’s multi-country analysis of children under 5 found increasing diarrhea risk with drought severity (45). Beyond overall risk, our findings suggest drought, like wet conditions, may shift the etiologic composition of diarrhea for admitted patients toward *V. cholerae*. The 4-week precipitation anomaly did not modify associations between drought and cholera, in contrast to findings that intense rainfall flushes fecal contamination into water sources and overwhelms sanitation infrastructure, increasing exposure to multiple enteric pathogens (29, 39, 46).

Our findings suggest WASHmobile could serve as a climate-resilient WASH intervention during drought conditions. During moderate drought significant effect modification was observed where the odds of handwashing with soap were lower in the standard arm, with no corresponding decline in the WASHmobile PICHA7 arm. This is consistent with the primary findings from this RCT, where WASHmobile significantly increased handwashing with soap throughout the 12-month follow-up (15). In this RCT, WASHmobile PICHA7 arm participants also had significantly lower *V. cholerae* serological responses than standard arm participants, providing strong evidence that the program has the potential to reduce cholera infection in this setting. Our similar RCT of WASHmobile in Bangladesh also found that WASHmobile significantly increased handwashing with soap throughout the 12 month follow-up and significantly reduced diarrhea (16). Previous studies show that drought may reduce handwashing by limiting water availability at handwashing stations, increasing time and effort to obtain water (47), or leading households to prioritize water for drinking and cooking over hygiene (48). Our findings support WASHmobile mobile messaging contributing to climate resilience by reinforcing handwashing with soap behaviors during extreme weather.

Our WASHmobile Anticipatory Action Framework builds on a growing number of anticipatory action frameworks for cholera response. In Cameroon, for example there is a Red Cross Early Action Protocol that combines climate risk data, weather forecasts, and community-based surveillance to create triggers that activate pre-arranged funding for water purification tablets, disinfection, and community risk communication (49). In the DRC, the United Nations Anticipatory Action Framework for cholera releases pre-arranged funding to support case management and WASH assistance when suspected cholera case thresholds are met (50). Our framework differs in three ways. First, its climate triggers are derived from associations between extreme weather, culture-confirmed cholera, and WASH behaviors observed in the same population, rather than from generic rainfall thresholds or relying only on case counts. Case counts are included in our framework as a complementary response trigger where ≥5 suspected cholera cases within 24 hours in a health area activates a targeted WASHmobile voice alert and chlorine e-vouchers for subscribers around the cholera case households based on our recent program evaluation (42). Second, its early actions are mechanism-specific WASH mHealth messages and chlorine e-vouchers which can be deployed quickly at low cost, rather than traditional WASH interventions and frequent in-person visits by health promoters. Third, it incorporates AI-enhanced forecasting systems such as Google’s Flood Hub and WeatherNext 3 to enhance the accuracy of flood-specific and weather-specific alerts (40, 41). Future studies are needed to investigate the implementation of our WASHmobile anticipatory action framework in different contexts globally.

This study has limitations. First, the pathogen specific type of diarrhea for cholera-negative patients was not ascertained, limiting our ability to identify how other pathogens responded to climatic exposures. Second, the study was conducted eastern DRC only, providing important context-specific evidence but limiting generalizability. The pre-defined climate triggers in our WASHmobile anticipatory action framework will need to tailored to the context where the framework will be implemented. Third, SPI-3 also reflects cumulative precipitation rather than flash flooding, a key hazard in the study area. Future studies should investigate the association between extreme weather and cholera over shorter time spans.

This study has several strengths. To our knowledge, it is the first study to evaluate how climatic conditions influence laboratory-confirmed cholera among admitted diarrhea patients using a test-negative design, providing pathogen-specific insights beyond analyses of all-cause diarrhea or suspected cholera cases. Our study is also among the first to assess climate resilience of a WASH intervention by examining whether climate modifies the intervention effects on WASH behaviors. These findings provide an empirical basis for pre-defined climate triggers and mechanism-specific early actions, which we translated into an anticipatory action framework for future a WASHmobile early warning system.

Drought and extreme wet conditions were associated with higher odds of cholera in eastern DRC. In our RCT, WASHmobile participants continue to have high handwashing with soap during drought conditions with significant declines observed among standard arm participants. This finding suggests WASHmobile could serve as a climate-resilient WASH intervention during drought. Study findings informed the WASHmobile anticipatory action framework where pre-defined climate triggers activate WASHmobile as an early warning system to reduce cholera outbreaks during extreme weather. Integrating climatic surveillance and forecasting with routine patient surveillance is critical for identifying periods of heightened cholera risk and informing timely, targeted WASH responses. Shifting cholera prevention from reactive outbreak response towards action based on climate forecasts and observed climate risks is critical to address cholera outbreaks from extreme weather events globally.

## Data Availability

All data produced in the present study are available upon reasonable request to the authors.

## ACKNOWLEDGEMENTS

The authors thank study participants. We would like to acknowledge research officers Pacifique Kitumaini, Gisèle Nsimire, Freddy Endelea, Feza Rugusha, Blessing Muderhwa Banywesize, Claude Lunyelunye, Emmanuel Buhendwa, Pascal Kitumaini Bujiriri, Jessy Tumusifu, and Brigitte Munyerenkana. The authors used Claude (Anthropic) to assist with grammar and clarity editing, checking internal consistency between the text and tables, coding review, and providing critical content review. All study design, data analysis, and interpretation were conducted by the authors, who reviewed and edited all AI-assisted suggestions and take full responsibility for the manuscript.

## REFERENCES

1. World Health Organization. Fact sheets: Climate change. 2023 [cited 2026 June 16]; Available from: https://www.who.int/news-room/fact-sheets/detail/climate-change-and-health

2. Borhani M, Nourani V, Wen H. Climate change and infectious diseases: breaking the poverty-disease-environment trap. Infectious Diseases of Poverty. 2026 2026/05/10;15(1):53.

3. Global Task Force for Cholera Control. Cholera Facts. [cited 2026 June 18]; Available from: https://www.gtfcc.org/about-cholera/cholera-facts/

4. World Meteorological Organization. El Niño set to become very strong, raising risks of extreme weather into 2027. Geneva, Switzerland: WMO; 2026.

5. Moore SM, Azman AS, Zaitchik BF, Mintz ED, Brunkard J, Legros D, et al. El Niño and the shifting geography of cholera in Africa. Proceedings of the National Academy of Sciences. 2017 2017/04/25;114(17):4436–41.

6. Dimitrova A, McElroy S, Levy M, Gershunov A, Benmarhnia T. Precipitation variability and risk of infectious disease in children under 5 years for 32 countries: a global analysis using Demographic and Health Survey data. Lancet Planet Health. 2022 Feb;6(2):e147–e55.

7. Doshi SM, Phillips MC, LaRocque RC, Duhaime A-C. Impact of climate change on diarrheal diseases: A scoping review. The Journal of Climate Change and Health. 2026 2026/03/01/;28:100612.

8. Nguyen AT, Grembi JA, Riviere M, Barratt Heitmann G, Hutson WD, Athni TS, et al. Influence of Temperature and Precipitation on the Effectiveness of Water, Sanitation, and Handwashing Interventions against Childhood Diarrheal Disease in Rural Bangladesh: A Reanalysis of the WASH Benefits Bangladesh Trial. Environ Health Perspect. 2024 Apr;132(4):47006.

9. Carlton EJ. Invited Perspective: Climate Changes the Effectiveness of Water, Sanitation, and Hygiene Interventions. Environ Health Perspect. 2024 Apr;132(4):41301.

10. Hubbard S, Wolf J, Oza HH, Arnold BF, Freeman MC, Levy K. Differential Effectiveness of Water, Sanitation, and Handwashing Interventions to Reduce Child Diarrhea in Dry and Rainy Seasons: A Systematic Review and Meta-Analysis of Intervention Trials. Environ Health Perspect. 2025 Feb;133(2):26001.

11. Ante-Testard PA, Rerolle F, Rahman M, Haque R, Das S, Parvez SM, et al. Seasonally Varying Effects of Improved Water, Sanitation, and Handwashing Interventions on Giardia Infection in Bangladesh. Open Forum Infect Dis. 2026 Aug;13(8):ofag430.

12. Ante-Testard PA, Rerolle F, Nguyen AT, Ashraf S, Parvez SM, Naser AM, et al. WASH interventions and child diarrhea at the interface of climate and socioeconomic position in Bangladesh. Nature Communications. 2024 2024/02/20;15(1):1556.

13. Alcayna T, Kellerhaus F, Tremblay L, Fletcher C, Goodermote R, Santos-Vega M, et al. Integrating anticipatory action in disease outbreak preparedness and response in the humanitarian sector. BMJ Glob Health. 2025 Jul 23;10(7).

14. WASHmobile Global. WASHmobile. 2026 [cited 2026 June 1]; Available from: https://www.washmobile.org/

15. George CM, Sanvura P, Bisimwa JC, Endres K, Namunesha A, Felicien W, et al. Effects of a Water, Sanitation, and Hygiene Program on Diarrhea, Cholera, and Child Growth in the Democratic Republic of the Congo: A Cluster-Randomized Controlled Trial of the Preventative Intervention for Cholera for 7 Days (PICHA7 WASHmobile) Mobile Health Program. Clin Infect Dis. 2025 Nov 6;81(4):785–96.

16. George CM, Monira S, Zohura F, Thomas ED, Hasan MT, Parvin T, et al. Effects of a Water, Sanitation, and Hygiene Mobile Health Program on Diarrhea and Child Growth in Bangladesh: A Cluster-randomized Controlled Trial of the Cholera Hospital-based Intervention for 7 Days (CHoBI7) Mobile Health Program. Clin Infect Dis. 2021 Nov 2;73(9):e2560-e8.

17. George CM, Namunesha A, Endres K, Felicien W, Sanvura P, Bisimwa JC, et al. Epidemiologic and Genomic Surveillance of Vibrio cholerae and Effectiveness of Single-Dose Oral Cholera Vaccine, Democratic Republic of the Congo. Emerg Infect Dis. 2025 Feb;31(2):288–97.

18. World Health Organization. Guidelines for drinking-water quality: fourth edition incorporating the first and second addenda. Geneva, Switzerland: World Health Organization; 2022.

19. Funk C, Peterson P, Landsfeld M, Pedreros D, Verdin J, Shukla S, et al. The climate hazards infrared precipitation with stations--a new environmental record for monitoring extremes. Sci Data. 2015 Dec 8;2:150066.

20. Armando CJ, Rocklöv J, Sidat M, Tozan Y, Mavume AF, Bunker A, et al. Spatial-temporal analysis of climate and socioeconomic conditions on cholera incidence in Mozambique from 2000 to 2018: an ecological longitudinal retrospective study. BMJ Open. 2024 Aug 19;14(8):e082503.

21. Alemayehu B, Ayele BT, Melak F, Ambelu A. Exploring the association between childhood diarrhea and meteorological factors in Southwestern Ethiopia. Sci Total Environ. 2020 Nov 1;741:140189.

22. Bradatan C, Dennis JA, Flores-Yeffal N, Swain S. Child health, household environment, temperature and rainfall anomalies in Honduras: a socio-climate data linked analysis. Environ Health. 2020 Jan 28;19(1):10.

23. Brintz BJ, Colston JM, Ahmed SM, Chao DL, Zaitchik BF, Leung DT. Assessment and comparison of model estimated and directly observed weather data for prediction of diarrhoea aetiology. Epidemiol Infect. 2024 Oct 9;152:e122.

24. Hajat S, Gampe D, Sarsour A, Abuzerr S. Climate Change and Diarrhoeal Disease Burdens in the Gaza Strip, Palestine: Health Impacts of 1.5 °C and 2 °C Global Warming Scenarios. Int J Environ Res Public Health. 2022 Apr 18;19(8).

25. Hossain N, Madaniyazi L, Ng CFS, Nasrin D, Seposo XT, Chua PLC, et al. Short-term associations of diarrhoeal diseases in children with temperature and precipitation in seven low- and middle-income countries from Sub-Saharan Africa and South Asia in the Global Enteric Multicenter Study. PLoS Negl Trop Dis. 2024 Oct;18(10):e0011834.

26. Huq A, Sack RB, Nizam A, Longini IM, Nair GB, Ali A, et al. Critical factors influencing the occurrence of Vibrio cholerae in the environment of Bangladesh. Appl Environ Microbiol. 2005 Aug;71(8):4645–54.

27. Wibawa BSS, Maharani AT, Andhikaputra G, Putri MSA, Iswara AP, Sapkota A, et al. Effects of Ambient Temperature, Relative Humidity, and Precipitation on Diarrhea Incidence in Surabaya. Int J Environ Res Public Health. 2023 Jan 28;20(3).

28. Andhikaputra G, Sapkota A, Lin YK, Chan TC, Gao C, Deng LW, et al. The impact of temperature and precipitation on all-infectious-, bacterial-, and viral-diarrheal disease in Taiwan. Sci Total Environ. 2023 Mar 1;862:160850.

29. Deshpande A, Chang HH, Levy K. Heavy Rainfall Events and Diarrheal Diseases: The Role of Urban-Rural Geography. Am J Trop Med Hyg. 2020 Sep;103(3):1043–9.

30. Haque F, Lampe FC, Hajat S, Stavrianaki K, Hasan SMT, Faruque ASG, et al. Impacts of climate change on diarrhoeal disease hospitalisations: How does the global warming targets of 1.5-2°C affect Dhaka, Bangladesh? PLoS Negl Trop Dis. 2024 Sep;18(9):e0012139.

31. Mertens A, Balakrishnan K, Ramaswamy P, Rajkumar P, Ramaprabha P, Durairaj N, et al. Associations between High Temperature, Heavy Rainfall, and Diarrhea among Young Children in Rural Tamil Nadu, India: A Prospective Cohort Study. Environ Health Perspect. 2019 Apr;127(4):47004.

32. Niven CG, Islam M, Nguyen A, Grembi JA, Mertens A, Pickering AJ, et al. Associations between weather extremes and faecal contamination along pathogen transmission pathways in rural Bangladeshi households: a prospective observational study. Lancet Planet Health. 2025 Jan;9(1):e5–e13.

33. Powers JE, Mureithi M, Mboya J, Campolo J, Swarthout JM, Pajka J, et al. Effects of High Temperature and Heavy Precipitation on Drinking Water Quality and Child Hand Contamination Levels in Rural Kenya. Environ Sci Technol. 2023 May 2;57(17):6975–88.

34. McKee TBD, N. J.; Kleist, J. The relationship of drought frequency and duration to time scales. Proceedings of the 8th Conference on Applied Climatology; 1993; Anaheim, CA, USA: American Meteorological Society; 1993. p. 179–83.

35. Beguería SV-S, Sergio M.; Angulo, Marta. SPEI: Calculation of the Standardized Precipitation-Evapotranspiration Index. 1.8.1 ed: Comprehensive R Archive Network (CRAN); 2023.

36. Yu W, Guo M, Li G, Li W, Cao Y, Hu K, et al. Compound drought and heatwave events and childhood diarrhea: current impacts and future risk in low- and middle-income countries. Environ Int. 2025 Sep;203:109792.

37. Keusch GT, Walker CF, Das JK. Diarrheal Diseases. In: Black RE, Laxminarayan R, Temmerman M, editors. Reproductive, Maternal, Newborn, and Child Health: Disease Control Priorities, Third Edition (Volume 2). Washington (DC): The International Bank for Reconstruction and Development / The World Bank; 2016.

38. WHO/UNICEF. The JMP service ladder for drinking water. 2026 [cited 2026 June 15]; Available from: https://washdata.org/topics/drinking-water

39. Kraay ANM, Man O, Levy MC, Levy K, Ionides E, Eisenberg JNS. Understanding the Impact of Rainfall on Diarrhea: Testing the Concentration-Dilution Hypothesis Using a Systematic Review and Meta-Analysis. Environ Health Perspect. 2020 Dec;128(12):126001.

40. Rasp S, Babenko B, Masters D, El-Kadi A, Merchant S, Shalev G, et al. WeatherNext 3: Increasing resolution and performance of global weather models with raw observations. arXiv. 2026.

41. Nearing G, Cohen D, Dube V, Gauch M, Gilon O, Harrigan S, et al. Global prediction of extreme floods in ungauged watersheds. Nature. 2024 Mar;627(8004):559–63.

42. George CM, Bisimwa JC, Endres K, Bengehya J, Kulondwa JC, Boroto R, et al. Program Evaluation of the WASHmobile PICHA7 Mobile Health and Chlorine E-Voucher Program in the Democratic Republic of the Congo. Trop Med Int Health. 2026 Jun;31(6):716–23.

43. Levy K, Woster AP, Goldstein RS, Carlton EJ. Untangling the Impacts of Climate Change on Waterborne Diseases: a Systematic Review of Relationships between Diarrheal Diseases and Temperature, Rainfall, Flooding, and Drought. Environmental Science & Technology. 2016 2016/05/17;50(10):4905–22.

44. Charnley GEC, Kelman I, Murray KA. Drought-related cholera outbreaks in Africa and the implications for climate change: a narrative review. Pathog Glob Health. 2022 Feb;116(1):3–12.

45. Wang P, Asare E, Pitzer VE, Dubrow R, Chen K. Associations between long-term drought and diarrhea among children under five in low- and middle-income countries. Nat Commun. 2022 Jun 30;13(1):3661.

46. Wang P, Asare EO, Pitzer VE, Dubrow R, Chen K. Floods and Diarrhea Risk in Young Children in Low- and Middle-Income Countries. JAMA Pediatr. 2023 Nov 1;177(11):1206–14.

47. Pinchoff J, Dougherty L, Dadi C. Water and Handwashing in a Drought-Prone Region of Southern Niger: How Environment, Household Infrastructure, and Exposure to Social and Behavior Change Messages Interact. The American Journal of Tropical Medicine and Hygiene. 2023 01 Mar. 2023;108(3):536–42.

48. Stoler J, Miller JD, Brewis A, Freeman MC, Harris LM, Jepson W, et al. Household water insecurity will complicate the ongoing COVID-19 response: Evidence from 29 sites in 23 low- and middle-income countries. Int J Hyg Environ Health. 2021 May;234:113715.

49. International Federation of Red Cross and Red Crescent Societies. Early Action Protocol: Cameroon - Cholera (Summary). Geneva, Switzerland: International Federation of Red Cross and Red Crescent Societies; 2025 2025/03/05.

50. United Nations Office for the Coordination of Humanitarian Affairs. DR Congo: Anticipatory Action Framework: Cholera - 2025. New York, NY, USA: United Nations Office for the Coordination of Humanitarian Affairs; 2025.

