## Appendix for "Evidence-Based WASHmobile Anticipatory Action Framework to Reduce Cholera Outbreaks during Extreme Weather Events in the Democratic Republic of the Congo"

**Supplemental File 1. WASHmobile PICHA7 RCT Method Detail**

Study Enrollment and Eligibility

WASHmobile PICHA7 RCT enrollment required that diarrhea patients meet the following criteria: (1) admission to a healthcare facility with three or more loose stools within a 24-hour period; (2) provision of both a blood and stool sample within 24 hours of enrollment; (3) presence of a child under 5 years of age in the household; (4) intention to reside in Bukavu for a minimum of 12 months; (5) having resided at home for the three nights preceding hospitalization; (6) absence of a functioning household tap for water collection (e.g., informal settlement); and (7) access to at least one working mobile phone within the household. Households were enrolled at the cluster level once an eligible diarrhea patient agreed to participate in the WASHmobile PICHA7 program, with additional household members eligible for inclusion if they had shared a cooking pot and residence with the patient for the prior 3 days and intended to continue residing together for the following 12 months. Participants were followed over a 12-month period.

Intervention Design

The WASHmobile PICHA7 program is structured around five key behaviors: (1) preparation of soapy water, using either water combined with detergent powder or water combined with small pieces of bar soap; (2) handwashing with soap or ash following stool-related and food-related events; (3) storage of drinking water in a vessel with both a lid and a tap; (4) treatment of household drinking water with chlorine tablets; and (5) boiling household drinking water for treatment when chlorine is not available. Households assigned to the WASHmobile PICHA7 arm received weekly mHealth over the 12-month follow-up period through a combination of automated voice calls, SMS messages, and interactive voice response (IVR) quizzes, delivered via the engageSPARK platform and framed as coming from a doctor at the local healthcare facility. Each IVR quiz presents participants with a question on a specific WASH behavior along with two possible answers; participants respond by pressing 1 or 2 and then receive feedback indicating whether their answer was correct, along with an explanation of the correct behavior.

RCT Outcomes

The WHO guideline for drinking water quality of <1 colony-forming unit (CFU)/100 mL of *E. coli* in drinking water was used as the cutoff to define safe drinking water quality relative to microbial contamination (1). The WHO free chlorine guideline of >0.2 mg/L for drinking water was used as the cutoff for a safe minimum drinking water level of free chlorine (1). Five-hour structured observations were conducted in a random subset of 50 households per study arm at Day 7 and at Months 1, 3, 6, 9, and 12 after enrollment to assess handwashing at food- and stool-related events. Handwashing with soap was recorded at the following key events promoted in the WASHmobile PICHA7 program: (1) after using the toilet; (2) after cleaning a child's anus; (3) after removing a child's feces; (4) before eating; (5) before feeding a child; (6) before preparing food; and (7) before serving food.

1. World Health Organization. Guidelines for drinking-water quality: fourth edition incorporating the first and second addenda. Geneva, Switzerland: World Health Organization; 2022.

**Appendix Table 1. Prevalence of each outcome within SPI-3 categories.**

| A | Cholera patient | | | | | | | | |
|---|---|---|---|---|---|---|---|---|---|
| | All ages (N=1068) | | | <5 years (N=520) | | | ≥5 years (N=548) | | |
| | % | n | N | % | n | N | % | n | N |
| Extreme drought | 18.8% | 3 | 16 | 0.0% | 0 | 7 | 33.3% | 3 | 9 |
| Severe drought | 46.7% | 7 | 15 | 12.5% | 1 | 8 | 85.7% | 6 | 7 |
| Moderate drought | 43.5% | 30 | 69 | 24.0% | 6 | 25 | 54.5% | 24 | 44 |
| Near normal | 28.4% | 206 | 725 | 9.2% | 35 | 380 | 49.6% | 171 | 345 |
| Moderate wet | 19.7% | 23 | 117 | 7.2% | 5 | 69 | 37.5% | 18 | 48 |
| Severe wet | 42.6% | 20 | 47 | 8.3% | 1 | 12 | 54.3% | 19 | 35 |
| Extreme wet | 46.8% | 37 | 79 | 36.8% | 7 | 19 | 50.0% | 30 | 60 |

| B | Handwashing with soap - all key events | | | | | | | | |
|---|---|---|---|---|---|---|---|---|---|
| | Overall (N=3397) | | | Standard arm (N=1566) | | | WASHmobile PICHA7 arm (N=1831) | | |
| | % | n | N | % | n | N | % | n | N |
| Extreme drought | – | 0 | 0 | – | 0 | 0 | – | 0 | 0 |
| Severe drought | 0.0% | 0 | 4 | 0.0% | 0 | 4 | – | 0 | 0 |
| Moderate drought | 25.0% | 34 | 136 | 1.3% | 1 | 78 | 56.9% | 33 | 58 |
| Near normal | 27.4% | 643 | 2345 | 7.4% | 81 | 1096 | 45.0% | 562 | 1249 |
| Moderate wet | 33.5% | 110 | 328 | 14.0% | 19 | 136 | 47.4% | 91 | 192 |
| Severe wet | 25.0% | 16 | 64 | 3.4% | 1 | 29 | 42.9% | 15 | 35 |
| Extreme wet | 32.3% | 168 | 520 | 4.9% | 11 | 223 | 52.9% | 157 | 297 |

| C | Stored Drinking Water E. coli > 1 CFU/mL | | | | | | | | |
|---|---|---|---|---|---|---|---|---|---|
| | Overall (N=1093) | | | Standard arm (N=591) | | | WASHmobile PICHA7 arm (N=502) | | |
| | % | n | N | % | n | N | % | n | N |
| Extreme drought | – | 0 | 0 | – | 0 | 0 | – | 0 | 0 |
| Severe drought | 0.0% | 0 | 1 | – | 0 | 0 | 0.0% | 0 | 1 |
| Moderate drought | 40.6% | 13 | 32 | 58.8% | 10 | 17 | 20.0% | 3 | 15 |
| Near normal | 28.7% | 254 | 886 | 41.8% | 205 | 490 | 12.4% | 49 | 396 |
| Moderate wet | 22.0% | 20 | 91 | 28.8% | 15 | 52 | 12.8% | 5 | 39 |
| Severe wet | 0.0% | 0 | 14 | 0.0% | 0 | 7 | 0.0% | 0 | 7 |
| Extreme wet | 11.6% | 8 | 69 | 28.0% | 7 | 25 | 2.3% | 1 | 44 |

| D | Stored Drinking Water free chlorine > 0.2 mg/L | | | | | | | | |
|---|---|---|---|---|---|---|---|---|---|
| | Overall (N=1167) | | | Standard arm (N=629) | | | WASHmobile PICHA7 arm (N=538) | | |
| | % | n | N | % | n | N | % | n | N |
| Extreme drought | – | 0 | 0 | – | 0 | 0 | – | 0 | 0 |
| Severe drought | 100.0% | 1 | 1 | – | 0 | 0 | 100.0% | 1 | 1 |
| Moderate drought | 58.3% | 21 | 36 | 27.8% | 5 | 18 | 88.9% | 16 | 18 |
| Near normal | 51.4% | 490 | 953 | 27.8% | 149 | 536 | 79.9% | 341 | 427 |
| Moderate wet | 62.4% | 58 | 93 | 41.5% | 22 | 53 | 90.0% | 36 | 40 |
| Severe wet | 71.4% | 10 | 14 | 57.1% | 4 | 7 | 85.7% | 6 | 7 |
| Extreme wet | 68.6% | 48 | 70 | 28.0% | 7 | 25 | 91.1% | 41 | 45 |

| E | Source Water E. coli > 1 CFU/mL | | | Source Water free chlorine > 0.2 mg/L | | |
|---|---|---|---|---|---|---|
| | Overall (N=796) | | | Overall (N=853) | | |
| | % | n | N | % | n | N |
| Extreme drought | – | 0 | 0 | – | 0 | 0 |
| Severe drought | – | 0 | 0 | – | 0 | 0 |
| Moderate drought | 37.9% | 11 | 29 | 34.4% | 11 | 32 |
| Near normal | 27.7% | 181 | 654 | 34.8% | 244 | 701 |
| Moderate wet | 24.1% | 14 | 58 | 23.7% | 14 | 59 |
| Severe wet | 11.1% | 1 | 9 | 33.3% | 3 | 9 |
| Extreme wet | 15.2% | 7 | 46 | 17.3% | 9 | 52 |

n is the number of observations with the outcome, N is the number of observations in the SPI-3 category (Supplemental Table 1), and % is n/N. The total analysis sample is given in each column heading. Cholera patients were defined as diarrhea patients with a V. cholerae-positive stool sample. Panels: (A) Proportion of admitted diarrhea patients with a V. cholerae-positive sample, by age group. (B) Handwashing with soap at all key events. (C) Stored drinking water E. coli >1 CFU/mL. (D) Stored drinking water free chlorine >0.2 mg/L. (E) Source water E. coli and free chlorine. Panels B–D are shown overall and by study arm, and panel E overall only. Near normal is the reference category in all regression models.

**Appendix Table 2A. Associations between SPI-3 drought categories and cholera and WASH outcomes, with the 4-week precipitation anomaly as an effect modifier.**

| | Severe drought vs. Near normal | | | Moderate drought vs. Near normal | | |
|---|---|---|---|---|---|---|
| | OR | 95% CI | p-value | OR | 95% CI | p-value |
| *Cholera patient* | | | | | | |
| All ages | | | | | | |
| 4 week precipitation anomaly | 1.00 | 0.82–1.22 | 0.970 | 1.00 | 0.82–1.22 | 0.970 |
| SPI-3 indicator | 5.58 | 0.53–59.06 | 0.153 | **2.39** | **1.33–4.28** | **0.004** |
| Interaction | 3.08 | 0.25–37.82 | 0.379 | 1.61 | 0.80–3.26 | 0.181 |
| ≥5 years | | | | | | |
| 4 week precipitation anomaly | 1.20 | 0.91–1.58 | 0.195 | 1.20 | 0.91–1.58 | 0.195 |
| SPI-3 indicator | 27.16 | 0.03–28800.00 | 0.353 | 1.63 | 0.74–3.59 | 0.226 |
| Interaction | 3.86 | 0.01–1540.00 | 0.658 | 1.55 | 0.58–4.13 | 0.378 |
| <5 years | | | | | | |
| 4 week precipitation anomaly | - | - | - | 0.84 | 0.55–1.28 | 0.410 |
| SPI-3 indicator | - | - | - | **3.65** | **1.28–10.40** | **0.015** |
| Interaction | - | - | - | 1.87 | 0.57–6.15 | 0.306 |

Each outcome is from a single model including SPI-3 category (near normal as reference), the 4-week precipitation anomaly (per 1 SD), and an SPI-3 drought category × 4-week precipitation anomaly interaction term. ORs are shown with 95% Wald CIs. Bold indicates $p < 0.05$, and "-" indicates the model could not be estimated due to sparse data. Cholera patients were defined as diarrhea patients with a V. cholerae-positive sample.

**Appendix Table 2B. Associations between SPI-3 drought categories and cholera, with models adjusted for vaccination status, gender, and age.**

| | Severe drought vs. Near Normal | | | Moderate drought vs. Near Normal | | |
|---|---|---|---|---|---|---|
| | OR | 95% CI | p-value | OR | 95% CI | p-value |
| *Cholera patient - adjusted for vaccination* | | | | | | |
| All ages | | | | | | |
| 4 week precipitation anomaly | 1.01 | 0.83–1.23 | 0.950 | 1.01 | 0.83–1.23 | 0.950 |
| SPI-3 indicator | 5.91 | 0.53–65.92 | 0.149 | **2.37** | **1.32–4.26** | **0.004** |
| Interaction | 3.27 | 0.25–42.38 | 0.365 | 1.62 | 0.80–3.28 | 0.176 |
| Over 5 | | | | | | |
| 4 week precipitation anomaly | 1.20 | 0.91–1.58 | 0.197 | 1.20 | 0.91–1.58 | 0.197 |
| SPI-3 indicator | 36.23 | 0.03–50000.00 | 0.331 | 1.65 | 0.74–3.65 | 0.219 |
| Interaction | 4.93 | 0.01–2520.00 | 0.616 | 1.59 | 0.59–4.27 | 0.354 |
| Under 5 | | | | | | |
| 4 week precipitation anomaly | - | - | - | 0.84 | 0.55–1.28 | 0.409 |
| SPI-3 indicator | - | - | - | **3.66** | **1.28–10.46** | **0.015** |
| Interaction | - | - | - | 1.87 | 0.57–6.16 | 0.305 |
| *Cholera patient - without vaccination* | | | | | | |
| All ages | | | | | | |
| 4 week precipitation anomaly | 1.01 | 0.83–1.24 | 0.885 | 1.01 | 0.83–1.24 | 0.885 |
| SPI-3 indicator | 3.74 | 0.39–36.15 | 0.255 | **2.29** | **1.26–4.14** | **0.006** |
| Interaction | 2.29 | 0.22–24.11 | 0.491 | 1.54 | 0.76–3.11 | 0.230 |
| Over 5 | | | | | | |
| 4 week precipitation anomaly | 1.20 | 0.90–1.59 | 0.210 | 1.20 | 0.90–1.59 | 0.210 |
| SPI-3 indicator | 14.24 | 0.03–7249.13 | 0.404 | 1.57 | 0.70–3.55 | 0.277 |
| Interaction | 2.44 | 0.01–475.48 | 0.740 | 1.49 | 0.55–4.00 | 0.431 |
| Under 5 | | | | | | |
| 4 week precipitation anomaly | - | - | - | 0.85 | 0.54–1.32 | 0.458 |
| SPI-3 indicator | - | - | - | **3.73** | **1.30–10.69** | **0.014** |
| Interaction | - | - | - | 1.85 | 0.56–6.13 | 0.317 |
| *Cholera patient - adjusted for gender* | | | | | | |
| All ages | | | | | | |
| 4 week precipitation anomaly | 1.00 | 0.82–1.22 | 0.973 | 1.00 | 0.82–1.22 | 0.973 |
| SPI-3 indicator | 5.58 | 0.53–58.67 | 0.152 | **2.39** | **1.33–4.28** | **0.004** |
| Interaction | 3.09 | 0.25–37.69 | 0.377 | 1.61 | 0.80–3.25 | 0.184 |
| Over 5 | | | | | | |
| 4 week precipitation anomaly | 1.20 | 0.91–1.58 | 0.198 | 1.20 | 0.91–1.58 | 0.198 |

| | Severe drought vs. Near Normal | | | Moderate drought vs. Near Normal | | |
|---|---|---|---|---|---|---|
| | OR | 95% CI | p-value | OR | 95% CI | p-value |
| SPI-3 indicator | 33.00 | 0.02–60400.00 | 0.362 | 1.57 | 0.71–3.45 | 0.265 |
| Interaction | 4.57 | 0.01–3010.00 | 0.646 | 1.63 | 0.61–4.36 | 0.326 |
| Under 5 | | | | | | |
| 4 week precipitation anomaly | - | - | - | 0.83 | 0.54–1.27 | 0.392 |
| SPI-3 indicator | - | - | - | **3.59** | **1.26–10.25** | **0.017** |
| Interaction | - | - | - | 1.83 | 0.55–6.06 | 0.322 |
| *Cholera patient - adjusted for age (years)* | | | | | | |
| All ages | | | | | | |
| 4 week precipitation anomaly | 1.01 | 0.83–1.24 | 0.883 | 1.01 | 0.83–1.24 | 0.883 |
| SPI-3 indicator | 5.81 | 0.48–70.80 | 0.168 | **2.10** | **1.16–3.80** | **0.014** |
| Interaction | 3.12 | 0.22–43.79 | 0.399 | 1.59 | 0.79–3.22 | 0.197 |
| Over 5 | | | | | | |
| 4 week precipitation anomaly | 1.23 | 0.93–1.63 | 0.156 | 1.23 | 0.93–1.63 | 0.156 |
| SPI-3 indicator | 33.48 | 0.02–58000.00 | 0.356 | 2.00 | 0.87–4.61 | 0.104 |
| Interaction | 4.12 | 0.01–2570.00 | 0.666 | 1.73 | 0.61–4.92 | 0.304 |
| Under 5 | | | | | | |
| 4 week precipitation anomaly | - | - | - | 0.84 | 0.52–1.37 | 0.496 |
| SPI-3 indicator | - | - | - | 3.24 | 0.95–11.08 | 0.061 |
| Interaction | - | - | - | 1.66 | 0.43–6.43 | 0.461 |

Each outcome is from a single model including SPI-3 category (near normal as reference), the 4-week precipitation anomaly (per 1 SD), and an SPI-3 drought category × 4-week precipitation anomaly interaction term. ORs are shown with 95% Wald CIs. Bold indicates $p < 0.05$, and "-" indicates the model could not be estimated due to sparse data. Without vaccination model only includes patients reporting no vaccination for cholera (N=1757). Cholera patients were defined as diarrhea patients with a V. cholerae-positive sample.

### Appendix Table 3A. Associations between SPI-3 categories and WASH outcomes, overall and by study arm.

| | | | | Moderate drought vs. near normal | | | Moderate wet vs. near normal | | | Severe wet vs. near normal | | | Extreme wet vs. near normal | | |
|---|---|---|---|---|---|---|---|---|---|---|---|---|---|---|---|
| | % | n | N | OR | 95% CI | p-value | OR | 95% CI | p-value | OR | 95% CI | p-value | OR | 95% CI | p-value |
| *Handwashing - all key events* | | | | | | | | | | | | | | | |
| Combined study arms | 28.3% | 939 | 3317 | 1.01 | 0.63–1.64 | 0.951 | 1.28 | 0.89–1.86 | 0.185 | 0.92 | 0.46–1.83 | 0.811 | 1.19 | 0.86–1.65 | 0.294 |
| WASHmobile PICHA7 arm | 46.6% | 828 | 1775 | 1.44 | 0.81–2.55 | 0.217 | 1.17 | 0.77–1.78 | 0.452 | 1.00 | 0.46–2.17 | 0.992 | 1.34 | 0.92–1.96 | 0.128 |
| Standard arm | 7.2% | 111 | 1542 | **0.16** | **0.03–0.81** | **0.027** | 1.74 | 0.91–3.32 | 0.094 | 0.77 | 0.21–2.85 | 0.689 | 0.68 | 0.31–1.47 | 0.325 |
| *Source water E. coli >1 CFU/mL* | | | | | | | | | | | | | | | |
| Combined study arms | 26.9% | 214 | 796 | 1.50 | 0.67–3.36 | 0.319 | 0.85 | 0.44–1.66 | 0.642 | 0.27 | 0.03–2.37 | 0.240 | **0.43** | **0.19–0.98** | **0.045** |
| *Source water chlorine* | | | | | | | | | | | | | | | |
| Combined study arms | 32.9% | 281 | 853 | 0.96 | 0.47–1.98 | 0.913 | 0.60 | 0.32–1.14 | 0.118 | 1.15 | 0.31–4.21 | 0.834 | **0.37** | **0.17–0.77** | **0.008** |
| *Stored water E. coli >1 CFU/mL* | | | | | | | | | | | | | | | |
| Combined study arms | 26.8% | 288 | 1075 | 1.93 | 0.86–4.35 | 0.113 | 0.65 | 0.36–1.16 | 0.142 | - | - | - | **0.41** | **0.20–0.86** | **0.018** |
| WASHmobile PICHA7 arm | 11.5% | 57 | 496 | 1.61 | 0.39–6.57 | 0.510 | 1.08 | 0.40–2.91 | 0.886 | - | - | - | 0.15 | 0.02–1.22 | 0.076 |
| Standard arm | 39.9% | 231 | 579 | 2.05 | 0.72–5.79 | 0.176 | **0.51** | **0.26–0.99** | **0.049** | - | - | - | 0.58 | 0.25–1.35 | 0.204 |
| *Stored Water free chlorine > 0.2 mg/L* | | | | | | | | | | | | | | | |
| Combined study arms | 54.3% | 623 | 1147 | 1.37 | 0.65–2.90 | 0.415 | **1.89** | **1.08–3.31** | **0.027** | 2.72 | 0.66–11.30 | 0.167 | 1.51 | 0.85–2.68 | 0.164 |
| WASHmobile PICHA7 arm | 82.3% | 437 | 531 | 2.08 | 0.58–7.46 | 0.260 | 2.40 | 0.67–8.61 | 0.179 | 1.73 | 0.33–9.04 | 0.517 | 2.72 | 0.66–11.30 | 0.167 |
| Standard arm | 30.2% | 186 | 616 | 1.13 | 0.38–3.34 | 0.827 | 1.69 | 0.88–3.25 | 0.113 | 3.65 | 0.79–16.89 | 0.097 | 0.99 | 0.39–2.52 | 0.986 |

ORs compare each SPI-3 category with near normal and are shown with 95% Wald CIs. %, n, and N are the outcome percentage, number of events, and analysis sample of each row's model. Bold indicates p < 0.05, and "-" indicates the model could not be estimated due to sparse data. Water quality models are adjusted for water source type (improved vs. unimproved per WHO JMP guidelines (38)). Handwashing and stored water models are adjusted for socioeconomic status (household income source, people per sleeping room, and concrete floor), and combined study arm models for study arm. Observations with incomplete socioeconomic data excluded.

**Appendix Table 3B. Associations between SPI categories and odds of cholera positivity of admitted diarrhea patients, with models adjusted for vaccination status, gender, and age.**

| | Extreme drought vs. near normal | | | Severe drought vs. near normal | | | Moderate drought vs. near normal | | | Moderate wet vs. near normal | | | Severe Wet vs. near normal | | | Extreme Wet vs. near normal | | |
|---|---|---|---|---|---|---|---|---|---|---|---|---|---|---|---|---|---|---|
| | OR | 95% CI | p-value | OR | 95% CI | p-value | OR | 95% CI | p-value | OR | 95% CI | p-value | OR | 95% CI | p-value | OR | 95% CI | p-value |
| *Cholera patient - adjusted for vaccination* | | | | | | | | | | | | | | | | | | |
| All ages | 0.57 | 0.16–2.03 | 0.387 | 2.21 | 0.79–6.18 | 0.130 | **1.92** | **1.16–3.18** | **0.011** | 0.62 | 0.38–1.00 | 0.052 | **1.85** | **1.01–3.37** | **0.045** | **2.24** | **1.40–3.59** | **0.001** |
| Over 5 | 0.50 | 0.12–2.02 | 0.329 | 6.40 | 0.76–54.08 | 0.088 | 1.22 | 0.65–2.29 | 0.540 | 0.62 | 0.33–1.15 | 0.129 | 1.20 | 0.59–2.41 | 0.615 | 1.02 | 0.59–1.77 | 0.933 |
| Under 5 | - | - | - | 1.41 | 0.17–11.83 | 0.750 | **3.12** | **1.17–8.36** | **0.023** | 0.77 | 0.29–2.04 | 0.599 | 0.90 | 0.11–7.18 | 0.920 | **5.73** | **2.11–15.57** | **0.001** |
| *Cholera patient - without vaccination* | | | | | | | | | | | | | | | | | | |
| All ages | 0.57 | 0.16–2.01 | 0.380 | 1.84 | 0.63–5.38 | 0.264 | **1.88** | **1.12–3.13** | **0.016** | 0.63 | 0.38–1.03 | 0.067 | 1.73 | 0.94–3.18 | 0.078 | **2.20** | **1.34–3.59** | **0.002** |
| Over 5 | 0.49 | 0.12–2.00 | 0.320 | 4.91 | 0.57–42.48 | 0.148 | 1.19 | 0.62–2.27 | 0.600 | 0.62 | 0.33–1.18 | 0.143 | 1.10 | 0.54–2.24 | 0.783 | 0.98 | 0.56–1.73 | 0.949 |
| Under 5 | - | - | - | 1.44 | 0.17–12.06 | 0.738 | **3.18** | **1.18–8.53** | **0.022** | 0.85 | 0.32–2.28 | 0.751 | 0.92 | 0.11–7.32 | 0.933 | **6.04** | **2.06–17.70** | **0.001** |
| *Cholera patient - adjusted for gender* | | | | | | | | | | | | | | | | | | |
| All ages | 0.58 | 0.16–2.06 | 0.399 | 2.20 | 0.79–6.15 | 0.132 | **1.94** | **1.17–3.21** | **0.010** | 0.62 | 0.38–1.00 | 0.051 | **1.86** | **1.02–3.39** | **0.043** | **2.22** | **1.39–3.55** | **0.001** |
| Over 5 | 0.52 | 0.13–2.14 | 0.367 | 6.16 | 0.73–52.03 | 0.095 | 1.15 | 0.61–2.17 | 0.667 | 0.58 | 0.31–1.09 | 0.090 | 1.23 | 0.61–2.49 | 0.560 | 0.98 | 0.57–1.71 | 0.953 |
| Under 5 | - | - | - | 1.39 | 0.17–11.65 | 0.760 | **3.10** | **1.16–8.28** | **0.024** | 0.77 | 0.29–2.04 | 0.596 | 0.88 | 0.11–7.01 | 0.901 | **5.67** | **2.09–15.37** | **0.001** |
| *Cholera patient - adjusted for age (years)* | | | | | | | | | | | | | | | | | | |
| All ages | 0.44 | 0.12–1.61 | 0.214 | 2.24 | 0.79–6.37 | 0.129 | **1.70** | **1.02–2.85** | **0.043** | 0.64 | 0.39–1.05 | 0.077 | 1.56 | 0.84–2.88 | 0.160 | **1.93** | **1.19–3.12** | **0.007** |
| Over 5 | 0.71 | 0.17–3.02 | 0.642 | 6.77 | 0.79–58.35 | 0.082 | 1.39 | 0.72–2.66 | 0.322 | 0.58 | 0.31–1.09 | 0.088 | 1.34 | 0.65–2.75 | 0.427 | 1.04 | 0.60–1.83 | 0.882 |
| Under 5 | - | - | - | 2.04 | 0.22–19.10 | 0.532 | 2.86 | 0.91–8.97 | 0.071 | 1.55 | 0.55–4.39 | 0.409 | 0.50 | 0.05–5.23 | 0.561 | **6.20** | **1.91–20.15** | **0.002** |

ORs compare each SPI-3 category with near normal and are shown with 95% Wald CIs. Bold indicates $p < 0.05$, and "-" indicates the model could not be estimated due to sparse data. Without vaccination model only includes patients reporting no vaccination for cholera (N=1757). Cholera patients were defined as diarrhea patients with a V. cholerae-positive sample.

**Appendix Table 4. Effect modification of associations between SPI-3 categories and WASH outcomes by study arm.**

| | Moderate drought vs. near normal | | | Moderate wet vs. near normal | | | Severe wet vs. near normal | | | Extreme wet vs. near normal | | |
|---|---|---|---|---|---|---|---|---|---|---|---|---|
| | OR | 95% CI | p-value | OR | 95% CI | p-value | OR | 95% CI | p-value | OR | 95% CI | p-value |
| *Handwashing - all key events* | | | | | | | | | | | | |
| SPI-3 indicator | **0.17** | **0.04–0.79** | **0.024** | 1.66 | 0.87–3.17 | 0.123 | 0.75 | 0.25–2.25 | 0.608 | 0.67 | 0.32–1.42 | 0.300 |
| Study arm | **8.34** | **5.76–12.08** | **<0.001** | **8.34** | **5.76–12.08** | **<0.001** | **8.34** | **5.76–12.08** | **<0.001** | **8.34** | **5.76–12.08** | **<0.001** |
| Interaction | **8.45** | **1.62–44.18** | **0.011** | 0.71 | 0.33–1.53 | 0.384 | 1.27 | 0.32–4.99 | 0.736 | 1.98 | 0.86–4.57 | 0.110 |
| *Stored water E. coli >1 CFU/mL* | | | | | | | | | | | | |
| SPI-3 indicator | 2.05 | 0.72–5.88 | 0.180 | 0.54 | 0.28–1.03 | 0.063 | - | - | - | 0.56 | 0.24–1.33 | 0.190 |
| Study arm | **0.20** | **0.14–0.30** | **<0.001** | **0.20** | **0.14–0.30** | **<0.001** | - | - | - | **0.20** | **0.14–0.30** | **<0.001** |
| Interaction | 0.84 | 0.15–4.66 | 0.846 | 1.89 | 0.57–6.20 | 0.296 | - | - | - | 0.29 | 0.03–2.58 | 0.269 |
| *Stored Water free chlorine > 0.2 mg/L* | | | | | | | | | | | | |
| SPI-3 indicator | 1.12 | 0.39–3.24 | 0.837 | 1.72 | 0.90–3.31 | 0.102 | 3.38 | 0.73–15.72 | 0.120 | 0.97 | 0.38–2.46 | 0.946 |
| Study arm | **10.61** | **7.40–15.22** | **<0.001** | **10.61** | **7.40–15.22** | **<0.001** | **10.61** | **7.40–15.22** | **<0.001** | **10.61** | **7.40–15.22** | **<0.001** |
| Interaction | 1.68 | 0.30–9.22 | 0.553 | 1.35 | 0.32–5.65 | 0.680 | 0.45 | 0.04–5.52 | 0.529 | 2.69 | 0.64–11.36 | 0.180 |

Each outcome is from one model including an SPI-3 category × study arm interaction. Within each category, the rows show: SPI-3 indicator: the OR for that category relative to near normal in the Standard arm; Study arm: the OR for the WASHmobile PICHA7 arm relative to the Standard arm; Interaction: the ratio by which the category's OR differs in the WASHmobile PICHA7 arm. ORs are shown with 95% Wald CIs. Bold indicates $p < 0.05$, and "-" indicates the model could not be estimated due to sparse data. Water quality models are adjusted for water source type (improved vs. unimproved per WHO JMP guidelines (38)). Handwashing and stored water models are adjusted for socioeconomic status (household income source, people per sleeping room, and concrete floor). Observations with incomplete socioeconomic data excluded.
